# Target-enhanced whole-genome sequencing (TE-WGS) shows clinical validity equivalent to commercially available targeted oncology panel

**DOI:** 10.1101/2023.12.20.23300156

**Authors:** Sangmoon Lee, Jin Roh, Jun Sung Park, Islam Oguz Tuncay, Wonchul Lee, Jung-Ah Kim, Brian Baek-Lok Oh, Jong-Yeon Shin, Jeong Seok Lee, Young Seok Ju, Ryul Kim, Seongyeol Park, Jaemo Koo, Hansol Park, Joonoh Lim, Erin Connolly-Strong, Tae-Hwan Kim, Yong Won Choi, Mi Sun Ahn, Hyun Woo Lee, Seokhwi Kim, Jang-Hee Kim, Minsuk Kwon

## Abstract

Cancer poses a significant global health challenge, with increasing incidence rates demanding precise genomic testing for individualized treatment strategies. Targeted-panel sequencing (TPS) has improved personalized oncology but often lacks comprehensive coverage of crucial cancer alterations. Whole-genome sequencing (WGS) addresses this gap, offering extensive genomic testing. This study demonstrates the potential medical application of WGS.

**Methods:** This study evaluates the power of target-enhanced WGS (TE-WGS), a clinical-grade WGS method sequencing both cancer and matched normal tissues. A cohort of forty-nine patients with various solid cancer types underwent both TE-WGS and TruSight Oncology 500 (TSO500), one of the mainstream TPS approaches currently used in the clinic.

**Results:** TE-WGS methods detected all the variants reported from TSO500 (100%, 498/498). A high correlation in the detection of variant allele fractions (VAF) was observed between the TE-WGS and TSO500 methodologies (r=0.977). Notably, within the pool of 498 variants commonly detected by both approaches, 223 variants (44.8%) were discerned within peripheral blood samples exclusively through the TE-WGS technique, suggesting their presence as constitutional variants inherent to the germline. Conversely, the remaining subset of 275 variants (55.2%) were not detected in peripheral blood using the TE-WGS method, signifying them as bona fide somatic variants. Further, TE-WGS provided accurate copy number profiles, fusion-genes, MSI- and homologous-recombination deficiency (HRD) scores, which were essential for clinical decision making.

**Conclusion:** TE-WGS proves to be a comprehensive approach in personalized oncology, matching the key biomarker detection capabilities of the established TSO500 panel. Additionally, TE-WGS uniquely identifies germline variants and genomic instability markers, offering additional clinical actions. Its adaptability and cost-effectiveness further underscore its clinical utility, making TE-WGS a valuable tool in personalized cancer treatment.

## INTRODUCTION

Cancer constitutes a substantial clinical burden worldwide, accompanied by the considerable cost of oncological therapies. In 2020, a staggering 19.3 million new cancer cases (18.1 million excluding nonmelanoma skin cancer) and nearly 10.0 million cancer-related deaths (9.9 million excluding nonmelanoma skin cancer) were reported globally[1]. In the United States alone, it was estimated that in 2023, there would be ∼2 million new cancer diagnoses and a projected 610K cancer-related deaths [2]. This escalating prevalence of cancer underscores an expanding, unmet demand for genomic testing—a pivotal tool empowering healthcare providers to adopt a more precise and tailored approach to patient care.

In the pursuit of personalized medicine within oncology, next-generation sequencing (NGS) has emerged as a valuable clinical asset. Recent decades of sequencing endeavors have unequivocally established cancer as predominantly driven by genetic factors[3]. The disease manifests through a complex interplay of inherited germline mutations and acquired somatic mutations, which can confer a survival advantage while evading immune surveillance [4]. Personalized medicine through harnessing genomic and molecular profiling technologies to customize treatments based on a patient’s specific tumor alterations, has demonstrated its potential[5-7]. It not only enhances survival rates and overall quality of life but also yields favorable economic outcomes when compared to single-gene tests [8,9]. Despite these promising advantages, the adoption of tumor profiling remains limited, with only a fraction of eligible patients benefiting from genomic testing [8,10]. Perhaps this underutilization is linked to the existing mainstream tests that often fall short of comprehensively covering all clinically relevant cancer-related alterations. Conventional targeted panel sequencing (TPS) approaches, for instance, require some prior level of diagnostic hypotheses driven by previous clinical and/or genomic data, which limits their clinical utility [4,11]. Additionally, due to validation cycles required for updating panel tests, they may struggle to address emerging biomarkers promptly [10]. These constraints underscore the pressing need for a comprehensive examination of a patient’s genome.

The rapid evolution of sequencing technologies, coupled with substantial cost reductions, has ushered in the era of comprehensive genomic profiling through whole-genome sequencing (WGS). WGS of tumor and matched normal tissues offers a comprehensive view of germline and somatic variants, including point mutations, copy number alterations, and structural rearrangements, delivering invaluable insights for precision oncology and personalized treatment strategies [12]. The comprehensive catalogues of mutations further allow more accurate quality control of genome sequences, and investigating patterns of mutations, known as “mutational signatures” [13]. As technology continues to advance and costs decline, the integration of WGS into routine comprehensive genomic profiling becomes increasingly indispensable to unlock the full potential of genomic medicine in cancer. Nonetheless, concerns have arisen regarding WGS’s sensitivity in detecting driver variants due to lower sequencing depth. Challenges also arise from low tumor cell fraction and suboptimal DNA quality from formalin-fixed paraffin-embedded (FFPE) tissue, potentially leading to the oversight of low allelic fraction variants by chance[14].

In response to these challenges, CancerVision introduces a forward-thinking approach—targeted enhanced WGS (TE-WGS). Here, on top of WGS backbone (40x), >500 top key biomarker genes are more deeply explored by cost-efficient enrichments using pools of WGS libraries (**Figure 1**). This unique amalgamation enhances confidence that even low-level expressions of critical biomarkers will not evade detection. This report presents the findings of TE-WGS CancerVision approach to cancer genomic profiling in comparison with TruSight^TM^ Oncology 500 (TSO500) in real-world clinical settings.

**Figure 1:**
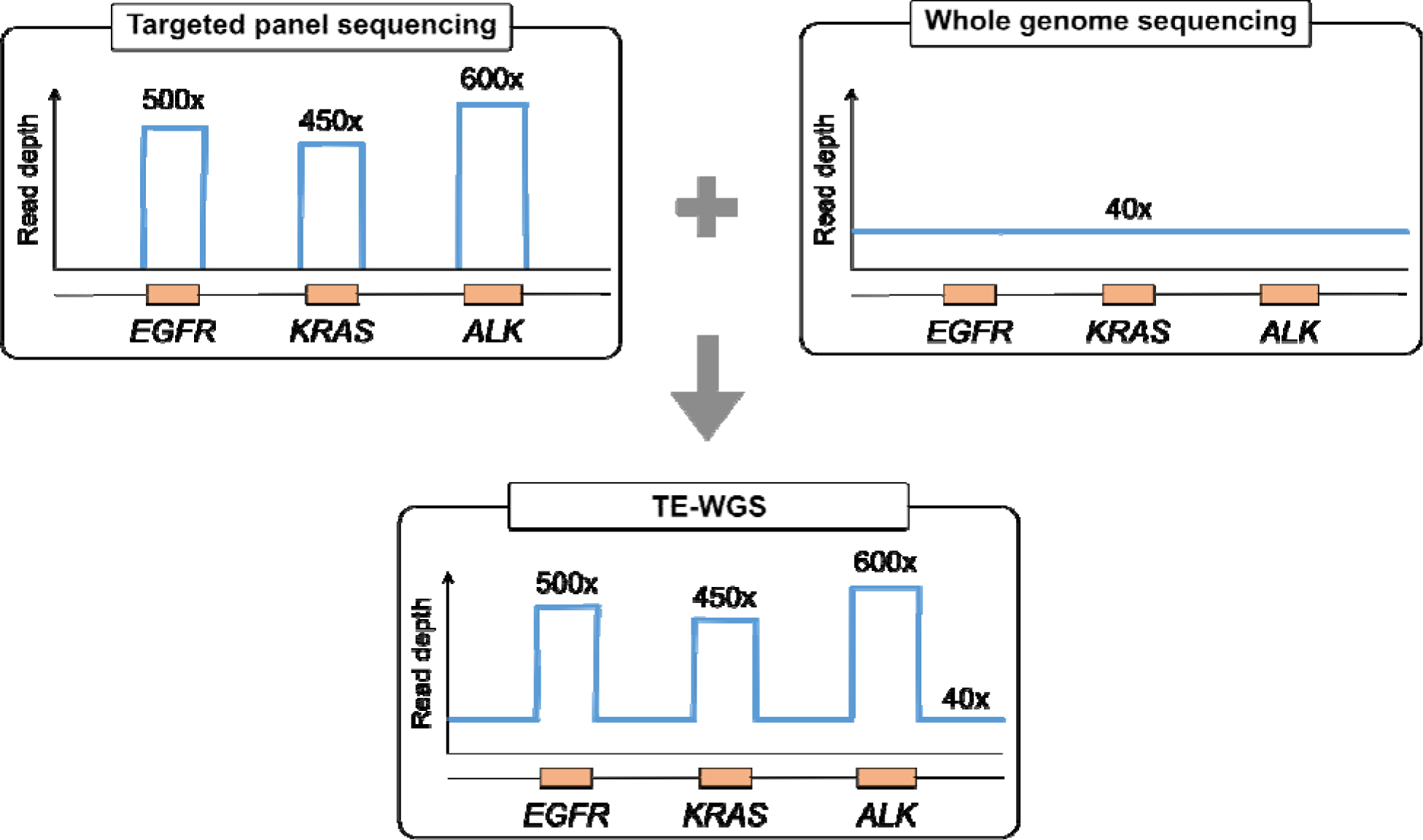
Overview of Target-Enhanced Whole-Genome Sequencing (TE-WGS). This cartoon depicts TE-WGS, which combines a 40x coverage WGS backbone with focused exploration of over 500 key biomarker genes. This is achieved through cost-efficient enrichment of pooled WGS libraries. The illustration highlights TE-WGS’s ability to detect low-level biomarker expressions, enhancing diagnostic precision in personalized oncology.

## METHODS

### Study Design

This prospective observational study included adult participants with diverse cancer types and stages, conducted at Ajou University Hospital from September 2022 to March 2023 (registered to Clinical Research Information Service, Number: KCT0007707). Each sample underwent diagnosis and tumor content assessment by a pathologist. All procedures received approval from the Institutional Review Board of Ajou University Hospital (AJOUIRB-SMP-2022-271). Routine cancer molecular profiling was performed on all patients using TruSight^TM^ Oncology 500 (TSO500) (Illumina Inc., San Diego, CA). Patients with residual tumor DNA after TSO500 testing provided peripheral blood samples for DNA extraction, enabling additional tumor-normal testing using Targeted Enhanced-Whole Genome Sequencing (TE-WGS) conducted by CancerVision, a proprietary Comprehensive Genomic Profiling (CGP) product developed by Genome Insight Inc., San Diego, CA. A total of 49 tumor-normal pairs were included in the study analysis (**Figure 2**).

**Figure 2:**
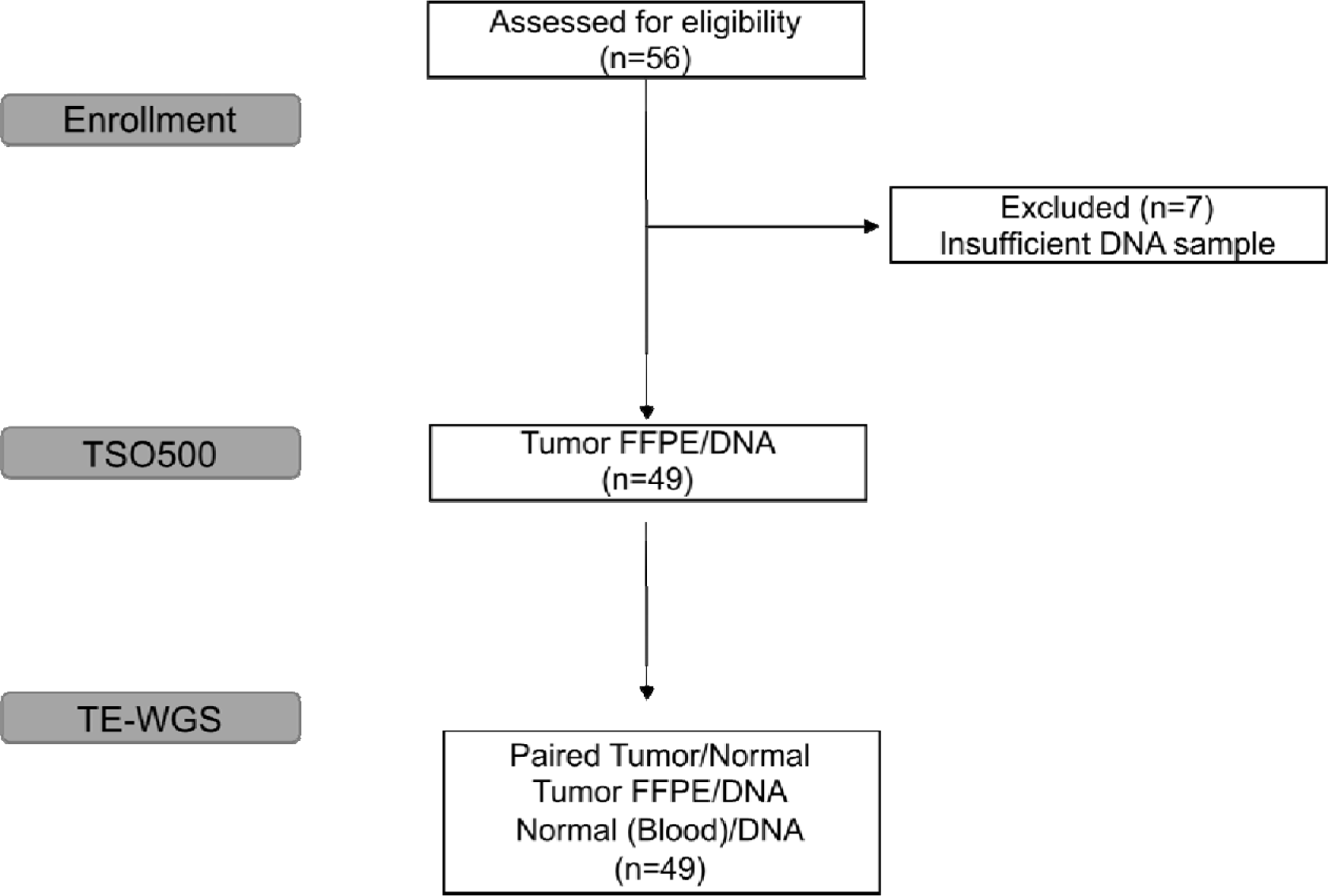
CONSORT diagram depicting patient enrollment and selection. Of the 56 patients enrolled in the study, only 49 met both the eligibility criteria and sample requirements, and thus, 49 patients were included in the analysis. TSO500 - TruSight Oncology 500; TE-WGS - Targeted Enhanced Whole Genome Sequencing.

### DNA Extraction

FFPE tumor DNA extraction was performed by the pathology laboratory at Ajou University Hospital using AllPrep DNA/RNA FFPE kit (Qiagen, Venlo, Netherlands). Peripheral blood DNAs were extracted using Allprep DNA/RNA kits (Qiagen) by Genome Insight Inc.

### Targeted Panel

Illumina’s TSO500 NGS library preparation adhered to the manufacturer’s instructions, with a DNA target size of 1.28Mb. Sequencing was executed on the NextSeq 550 platform (Illumina Inc.), and data analysis occurred via TruSight Oncology 500 Local App (Illumina Inc., pipeline version 2.2) on an on-premise server at Ajou University Hospital. Reported findings encompassed SNVs and INDELs with a Variant Allele Frequency (VAF) >= 5%, Copy Number Variations (CNVs) with >= 4 (gain) and <1 (loss), and translocations supported by >=5 translocation reads.

### Targeted-Enhanced Whole Genome Sequencing (TE-WGS)

Residual tumor DNA from TSO500 testing by Ajou University Hospital and newly extracted peripheral blood DNA from Genome insight Inc. were utilized to prepare paired tumor-normal sequencing libraries. Genome sequencing, analysis, and interpretation were conducted using the CancerVision system (Genome Insight Inc., San Diego, CA, USA). DNA libraries were created with TruSeq Nano Library Prep Kits (Illumina Inc.) and sequenced on the Illumina NovaSeq6000 platform (Illumina Inc.) with average depths of coverage of 40x for tumor samples and 20x for blood samples for WGS. For target enhancement, tumor DNA libraries underwent capture using xGen Custom Hybridization Probes from IDT, Inc. (Coralville, IA, USA) with a target size of 2.76 Mb. Sequencing was performed on the Illumina NovaSeq6000 platform with an average depth of coverage of 500x.

The obtained sequences were aligned to the human reference genome (GRCh38) using the BWA-MEM algorithm. PCR duplicates were removed using SAMBLASTER [15]. Initial germline mutation calling for small variants was performed using HaplotypeCaller2 [16] and Strelka2 [11], and somatic mutation for small variants was performed using Strelka2 [11] and Mutect2[17]. Structural variations were identified using Manta [18]. Identified germline and somatic variants were annotated using VEP [19] and manually inspected and curated using Genome Insight’s proprietary genome browser.

## RESULTS

### Demographics

A total of 56 patients diagnosed with solid tumors between September 2022 and March 2023 were initially enrolled in the study. However, seven patients were subsequently excluded from future analysis due to insufficient DNA samples. Thus, a final cohort of 49 patients with available tumor-normal pairs was included in the subsequent analysis (**Figure 2**). Among the participants, 31 (63%) were male, while the remaining 18 (37%) were female. The median age of the cohort was 59 years (range 22 – 86 years). The cohort encompassed a diverse array of tumor types, as detailed in Table 1. Briefly, these tumor types included breast (n=4), colorectal (n=7), pancreatic (n=11), stomach (n=8), and lung cancers (n=4).

**Table 1:** Demographics.

| Patient Characteristics |  | n (%) |
| --- | --- | --- |
| Sex |  |  |
|  | Male | 31(63) |
|  | Female | 18 (37) |
| Age in years |  |  |
|  | Mean | 58.8 |
|  | Range | 22-86 |
| Tumor Type |  |  |
|  | Colorectal adenocarcinoma | 7 (14) |
|  | Breast invasive ductal carcinoma | 4 (8) |
|  | Lung adenocarcinoma | 2 (4) |
|  | Lung squamous cell carcinoma | 2 (4) |
|  | Pancreatic adenocarcinoma | 11 (22) |
|  | Renal cell carcinoma | 3 (6) |
|  | Stomach adenocarcinoma | 8 (16) |
|  | Urothelial cancer | 3 (6) |
|  | Other | 9 (18) |

### Coverage depth

The average, mean depth of coverage for TSO500 was 830.1x (range 378.5-1035.1). In the TE-WGS approach, cancer whole-genome was sequenced to the mean depth of ∼45x (range 28.4-68.5) with an enrichment for target genes to 359.8x (range 84.4-896.5). Notably, 93.12% of the targeted regions achieved a sequence depth of 50x in the TE-WGS approach, whereas this value was 98.28% for TSO500. (**Figure 3A**)

**Figure 3:**
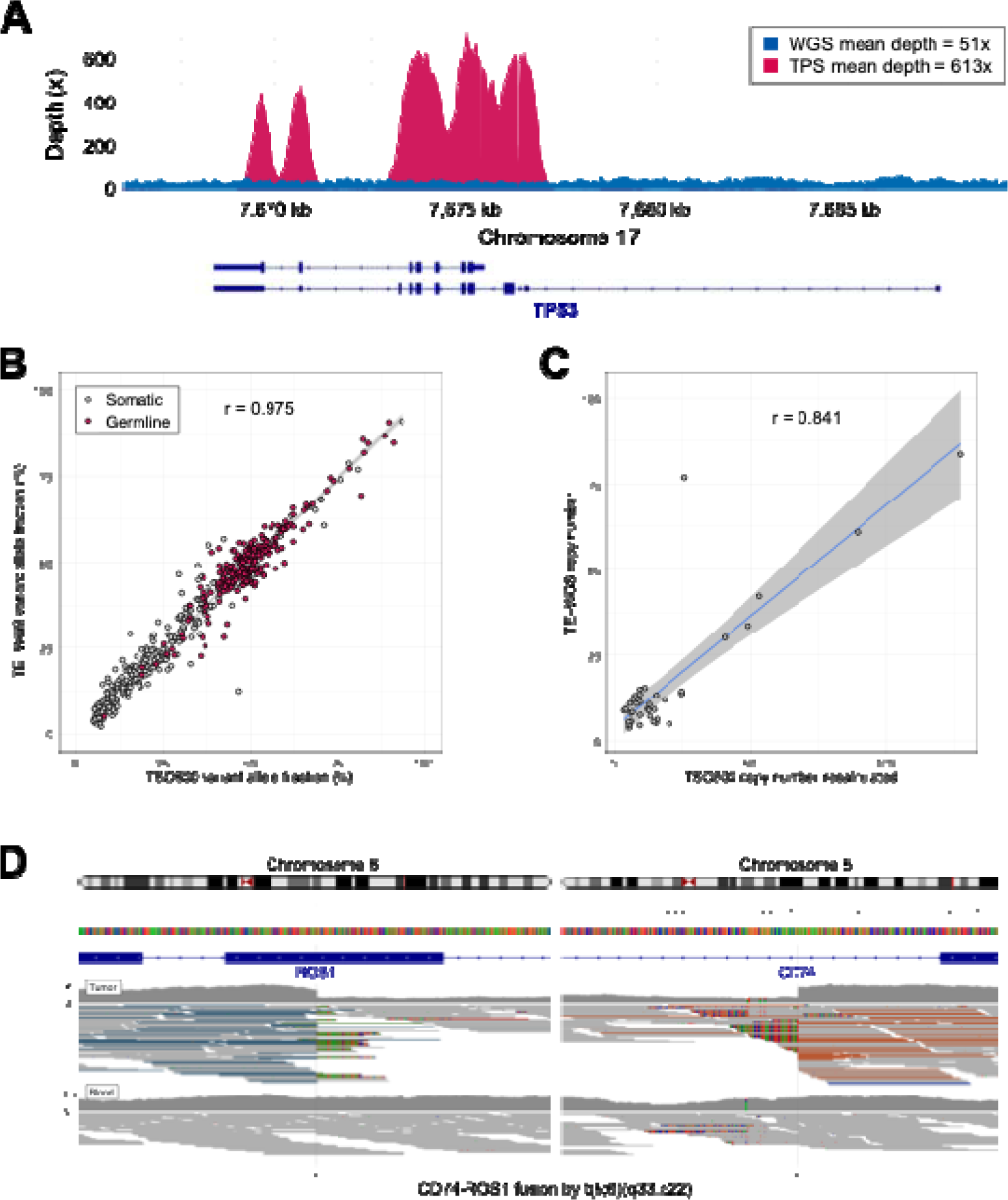
Variant comparison between TSO500 and TE-WGS. (A) Representative sequencing depth on TP53 locus for TE-WGS. In general, TE-WGS achieved a 50x sequence depth in 93.12% of targeted regions. (B) Complete correlation in variant allele fractions (VAF) detected between TSO500 and TE-WGS methodologies (r=0.975). Of the shared variants, TE-WGS determined 44.8% of the variant to be of germline origin (red color), while 55.2% are identified as somatic variants. (C) Concordance in gene amplification copy number between TSO500 and TE-WGS, after adjusting for tumor purity by TE-WGS (r = 0.842, p = 7.24e-11). All tier I and II CNVs from TSO500 were confirmed by TE-WGS. (D) CD74::ROS1 fusion in lung adenocarcinoma was identified by TE-WGS.

### Variants detection

In order to evaluate the TE-WGS performance in variant detection of clinically actionable driver genes, genomic findings from the TSO500 were first considered. In total, 498 unique variants were identified across all samples in TSO500, with a complete detection in the TE-WGS methods, accounting for the 100% sensitivity (498/498). The data demonstrated a high correlation of detection of variant allele fractions (VAF) between the TSO500 and TE-WGS methodologies (r=0.975) (**Figure 3B**).

Furthermore, among the pool of 498 commonly detected variants identified by both approaches, a notable proportion of 223 variants (44.8%) were discerned within peripheral blood samples through the TE-WGS technique, suggestive of constitutional variants inherent to the germline. Conversely, the remaining subset of 275 variants (55.2%) were not detected in peripheral blood through the TE-WGS method, signifying them as bona fide somatic variants of the tumor tissue. Within the tumor TE-WGS data, the VAFs exhibited a spectrum of 5.0-90.7% for constitutional variants and 2.1-91.0 % for somatic variants, with a substantial level of overlap. This underscores that the VAF may not be relied upon as a definitive criterion to distinguish germline and somatic variants (**Figure 3B**).

### Tumor Cell Fraction/Copy Number Analysis

The TSO500 assay does not employ an algorithmic process to calculate tumor cell fraction (TCF); rather, it incorporates estimations provided by pathologists into the final report. In contrast, the TE-WGS approach utilizes a computational algorithm that considers copy number variations and allelic balance changes. While there exists a correlation between TCF calculations using the TE-WGS method and pathologists’ estimates (r=0.44), a discernible discrepancy in estimation between the two methodologies is evident. This observed incongruity can be attributed to the well-documented tendency of pathologists to provide higher TCF estimations compared to those derived from the WGS-based approach [20]. (**Figure 4A**)

**Figure 4:**
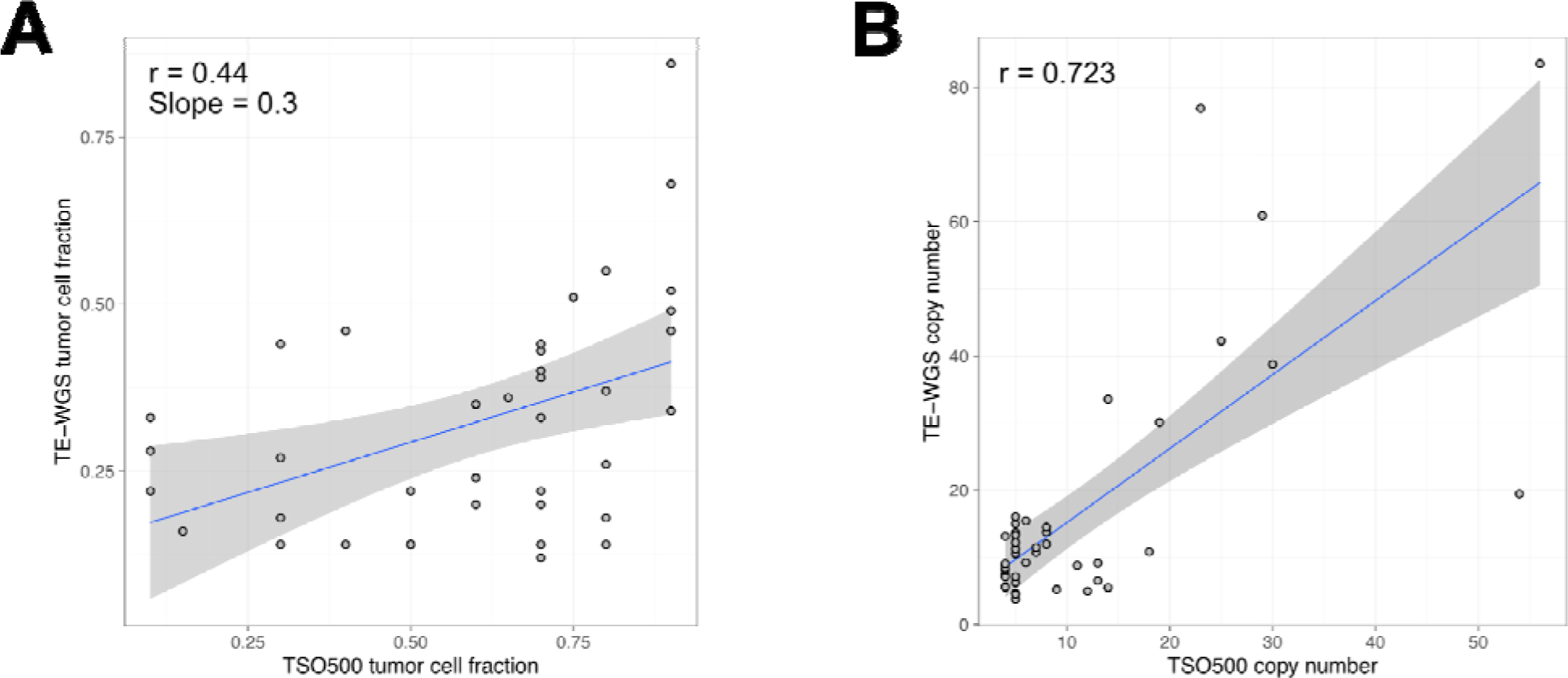
Tumor cell fraction (TCF) estimation (A) TSO500’s reliance on pathologists’ inputs and TE-WGS’s computational algorithm, leading to evidence discrepancies and a weak correlation (r=0.44) (B) Concordance in gene amplification copy number between TSO500 and TE-WGS, before adjusting for tumor purity by TE-WGS.

Copy number of gene amplifications reported by TSO500 was overall concordant to the TE-WGS (r = 0.723, p = 4.19e-08, **Figure 4B**). Given that tumor purity determined by pathologists is taken into account to calculate the gene copy number of TSO500, we recalculated the copy number using tumor purity more accurately determined by TE-WGS. As a result, the copy numbers between TSO500 and TE-WGS showed higher correlation (r = 0.842, p = 7.24e-11, **Figure 3C**). All the tier I and II CNVs from TSO500 were also detected by TE-WGS.

### Gene Fusions analysis

TSO500 identified two fusion events which were actionable by the CKB (Jackson Laboratory, Bar Harbor, ME). These fusion events were associated with an AMP/ASCO/CAP Consensus Recommendation level A or D[21]: CD74::ROS1 in lung adenocarcinoma and TMPRSS2::ERG in prostate cancer. TE-WGS approach reported both fusions from WGS data. (**Figure 3D**)

### Genomic Instability Analysis

Tumor Mutation Burden (TMB): The TSO500 assay estimates TMB by calculating the number of passing eligible variants and dividing it by the size of the targeted coding region covered by the panel. In contrast, the TE-WGS approach employs a germline subtraction method to determine the number of somatic alterations per million base pairs across the entire genome. A TMB value exceeding 10 mutations per megabase (Mb) is classified as ‘high’ (≥10mut/Mb)[22]. TMB scores calculated by TE-WGS exhibited a good correlation with those derived from TSO500 (r=0.89, **Figure 5A**). However, some instances of discordance were observed. Notably, six (12.2%) TSO500-tested tumors classified as TMB-high were categorized as TMB-low (<10 mutations/Mb) by the TE-WGS approach (**Figure 5B**)

**Figure 5:**
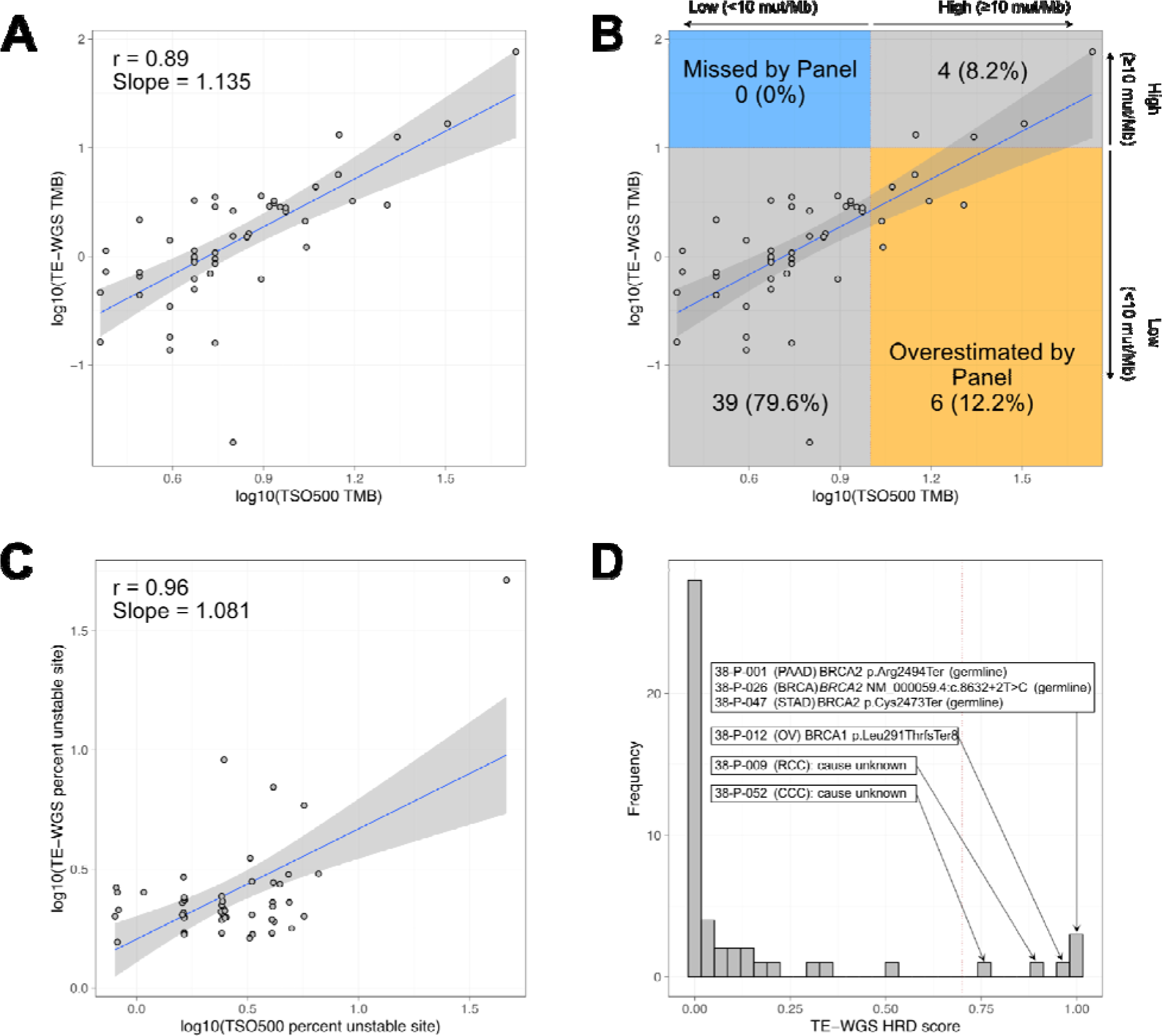
Comparison of genomic instability biomarkers. (A) Comparison of Tumor Mutation Burden (TMB) assessment between TSO500 and TE-WGS reveals a good correlation (r=0.89), though some discrepancies were observed in TMB-high classification. (B) 12.2% of tumors classified as TMB-high by TSO500 were deemed TMB-low by TE-WGS. (C) Evaluation of microsatellite instability (MSI) showed strong concordance between TSO500 and TE-WGS (r=0.96); most tumors were MSI stable, with one notable exception. (D) TE-WGS’s method for assessing homologous recombination deficiency (HRD) effectively distinguished HR-proficient from HR-deficient tumors, revealing germline variants in HRD-related genes not identified by TSO500.

Microsatellite instability (MSI): TSO500 relies on stitched reads obtained from tumor samples to determine the MSI status. Unstable microsatellite sites are detected by assessing the shift in the length of a microsatellite site for a tumor sample against a set of normal baseline samples. In contrast, TE-WGS employs a proprietary algorithm that incorporates germline sequence subtraction and detects somatic microsatellite changes for MSI estimation. There was a strong correlation between these two methodologies in MSI scoring (r=0.96) (**Figure 5C**). The majority of tumors in the cohort were MSI stable. However, there was one ureter cancer that was identified by TSO500 as MSI-high, and confirmed to be MSI-high by TE-WGS. Further, we detected *MSH2* (NM_000251.3:c.1043del, p.Gln348ArgfsTer9) somatic mutation which likely underlies the MSI-high phenotype in the case.

Homologous recombination deficiency (HRD): TSO500 does not include HRD evaluation, Illumina offers an optional add-on Kit for HRD assessment. However, this supplementary kit was not integrated into our study design, thus preventing a direct head-to-head comparison of HRD scoring between the two methodologies. The TE-WGS approach employed a proprietary algorithm dedicated to HRD scoring. Notably, this method effectively discriminated between HR-proficient and HR-deficient tumors (**Figure 5D**). A majority of HR-deficient tumors exhibited germline variants in HRD-related genes, including *BRCA1* and *BRCA2*. While TSO500 did detect *BRCA1/2* variants in cases identified by TE-WGS, it did not provide informative HRD assessment.

### Mutational Signature

WGS is an unbiased approach to profiling mutational signatures [23]. Mutational signature analysis identified tumor samples with homologous recombination deficiency (HRD), characterized by enrichment of single base substitution (SBS) signature 3 and small insertions and deletions (ID) signature 6 (**Figure 6**). Mutational signature analysis also identified tumor samples with APOBEC (apolipoprotein B mRNA-editing catalytic polypeptide) mutations, characterized by enrichment of SBS2 and SBS13 signatures (**Figure 6**). Identification of the HRD mutational signature suggests that the mutational process is caused by HRD, even if the related genes, such as *BRCA1/2*, are not found.

**Figure 6:**
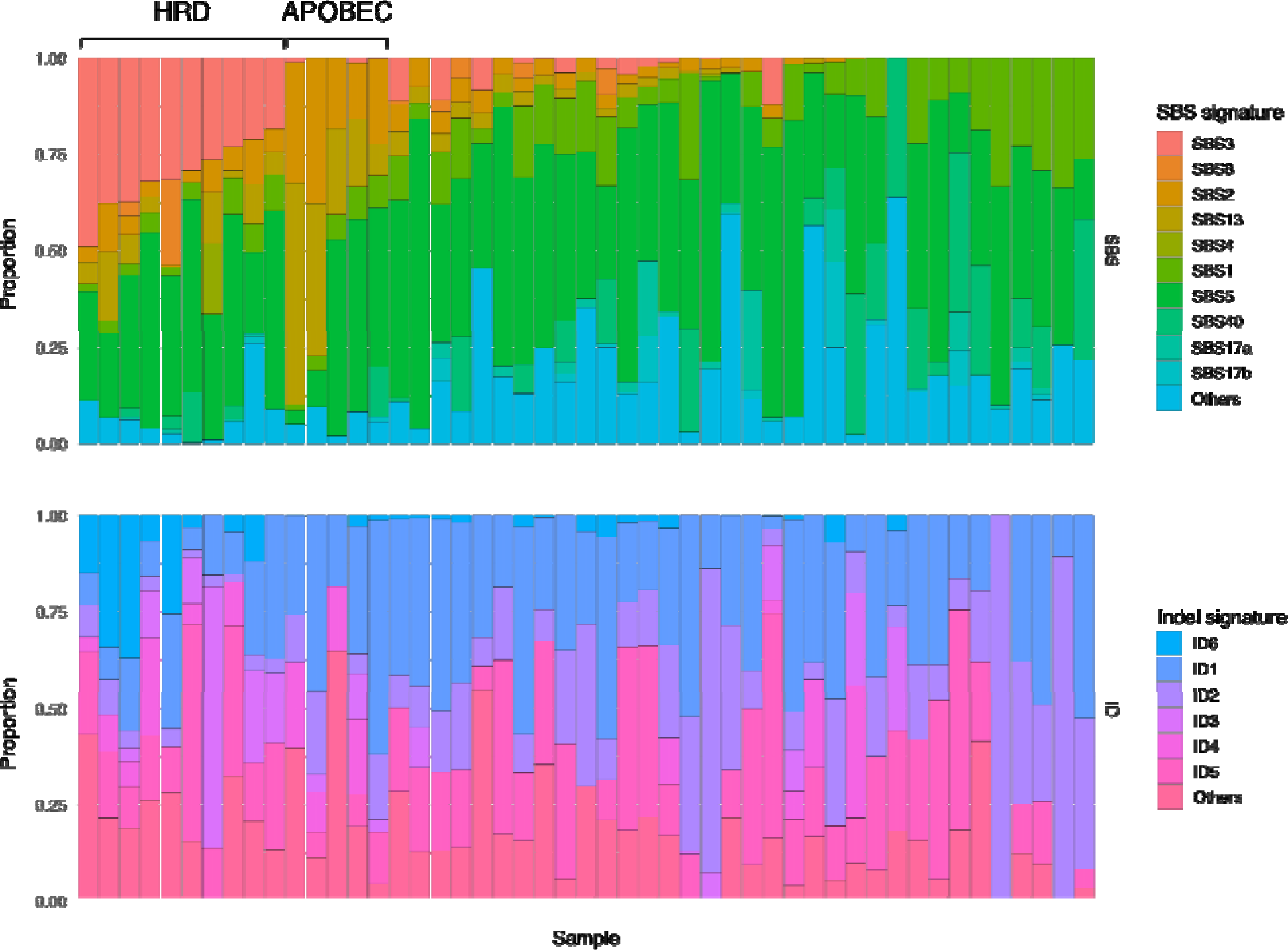
Mutational signature analysis highlighting tumor samples exhibiting homologous recombination deficiency (HRD), as evidenced by the enrichment of single base substitution (SBS) signature 3 and small insertions and deletions (ID) signature 6, and tumors bearing APOBEC mutations, characterized by the enrichment of SBS2 and SBS13 signatures. The presence of the HRD mutational signature suggests an underlying HRD mutational process, even in the absence of related gene mutations like *BRCA1/2*.

## DISCUSSION

Cancer remains a formidable challenge in modern medicine, despite remarkable advances in treatment modalities such as targeted therapies and immunotherapies. These breakthroughs have ushered in the era of precision medicine, substantially improving survival rates across various cancer types [20]. However, the clinical application of these therapies hinges on the accurate identification of actionable genomic alterations. Next Generation Sequencing (NGS) technologies have demonstrated their clinical utility by enabling the identification of such alterations, facilitating informed decisions regarding approved therapies, clinical trial participation, or the management of rare actionable mutations [24,25].

Nonetheless, a crucial limitation of fixed-panel NGS assays like TruSight Oncology 500 (TSO500) has hindered their ability to keep pace with new discoveries. These panels are inherently constrained by the genomic alterations known at the time of their design. As new biomarkers emerge, these tests quickly become outdated, depriving patients of access to the latest insights that could guide their care. Moreover, the development and validation of updated panels introduce a lag that impedes the timely adoption of emerging biomarkers [11,26,27], necessitating novel approaches.

Targeted-Enhanced Whole Genome Sequencing (TE-WGS) offers an intriguing solution to this predicament. By combining a deep-read panel with a Whole Genome Sequencing (WGS) framework, TE-WGS provides the most comprehensive view of the genomic landscape, unburdened by the constraints of predefined gene panels. This adaptability allows for swift algorithm updates without the need for creating and validating entirely new tests. Additionally, the WGS approach inherently possesses the ability to retrospectively identify emerging biomarkers in previously sequenced patient data, ensuring that patients benefit from the latest discoveries, even after their initial testing.

The clinical utility of targeted genomic panels, encompassing both germline susceptibility and known “actionable” somatic mutations (including TSO500), has been well-documented [27-33]. For some cancer types, targeted panel sequencing is becoming standard practice [31]. This study demonstrated a robust correlation between TSO500 and TE-WGS in detecting single nucleotide variants (SNVs), insertions/deletions (indels), and copy number variations (CNVs). This correlation implies that the clinical utility demonstrated by TSO500 can also be applied to the actionable key biomarker findings reported by the TE-WGS (CancerVision) test. Furthermore, TE-WGS extends the actionability seen with TSO500 by reporting the origin (somatic or germline) of actionable biomarkers. Distinguishing between these categories allows for more informed treatment decisions, potentially reducing the risk of exposure to ineffective therapeutic approaches.

In recent years, genomic instability markers have gained attention with the advent of immunotherapy. Tumor Mutational Burden (TMB) has emerged as a predictive biomarker for immunogenic neoantigens [6,7]. However, studies have suggested that TMB calculations using tumor-only assays can yield falsely elevated results compared to those determined by germline subtraction methodologies [34]. The TE-WGS approach employs a proprietary algorithm informed by germline subtraction and whole genome analysis. While this study showed a good correlation between TSO500 and TE-WGS approaches, instances of discordance were observed, indicating that TE-WGS methodology may have the potential to reduce exposure to ineffective therapeutic approaches.

Beyond the classical clinical utility of targeted panels, the TE-WGS approach can detect somatic variants arising from specific mutagenic mechanisms or signatures. Each mutagenic source produces a characteristic mutational pattern, and unsupervised learning techniques can decipher signatures associated with distinct etiologies. These signatures, often referred to as “genomic scars,” represent lasting and detectable evidence of environmental or internal genomic damage [13,35]. The most notable and actionable mutational signature currently is the Homologous Recombination Deficiency (HRD) signature. Tumors with mutations in *BRCA1/2* are deficient in the Homologous Recombination Repair (HRR) process, and they exhibit promising responses to PARP inhibitors[36]. The TE-WGS approach allows for bioinformatic integration of known mutational signatures (COSMIC mutational signatures [37]) in a clinical setting, eliminating the need for additional testing for each mutational signature.

Economic studies have demonstrated that using NGS panel testing in patients is associated with cost savings for both CMS and commercial payers compared to standard limited gene testing [26,38,39]. However, the need for multiple panel tests, including somatic and germline testing, represents an area of inefficient spend for both CMS and commercial payers. The TE-WGS approach offers a cost-effective solution for complete genomic profiling, reducing the time to reporting as germline, somatic, genomic instability, and mutation signature findings are obtained from one test. This approach not only allows for cost-effectiveness but also streamlines the diagnostic process.

In conclusion, Targeted-Enhanced Whole Genome Sequencing (TE-WGS) emerges as a promising and potent approach in the pursuit of personalized medicine in oncology. It effectively addresses the need for comprehensive profiling while remaining cost-effective, and adaptable to emerging biomarkers.

## Data Availability

All data produced in the present study are available upon reasonable request to the authors.

## Acknowledgments

This work was supported by the National Research Foundation of Korea (NRF) grant funded by the Korea government (MSIT) (No. NRF-2022R1F1A1074910). This research was supported by a grant of the Korea Health Technology R&D Project through the Korea Health Industry Development Institute (KHIDI), funded by the Ministry of Health & Welfare, Republic of Korea (HR22C1734, HI23C1589).

## REFERENCES

[1] Sung H, Ferlay J, Siegel RL, et al. Global Cancer Statistics 2020: GLOBOCAN Estimates of Incidence and Mortality Worldwide for 36 Cancers in 185 Countries. CA Cancer J Clin. 2021 May;71(3):209-249.

[2] Siegel RL, Miller KD, Wagle NS, Jemal A. Cancer statistics, 2023. CA Cancer J Clin. 2023 Jan;73(1):17-48.

[3] Lee H, Palm J, Grimes SM, Ji HP. The Cancer Genome Atlas Clinical Explorer: a web and mobile interface for identifying clinical-genomic driver associations. Genome Med. 2015 Oct 27;7:112.

[4] Rosenquist R, Cuppen E, Buettner R, et al. Clinical utility of whole-genome sequencing in precision oncology. Semin Cancer Biol. 2022 Sep;84:32–39.

[5] Paik S, Shak S, Tang G, et al. A multigene assay to predict recurrence of tamoxifen-treated, node-negative breast cancer. N Engl J Med. 2004 Dec 30;351(27):2817–26.

[6] Marabelle A, Fakih M, Lopez J, et al. Association of tumour mutational burden with outcomes in patients with advanced solid tumours treated with pembrolizumab: prospective biomarker analysis of the multicohort, open-label, phase 2 KEYNOTE-158 study. Lancet Oncol. 2020 Oct;21(10):1353–1365.

[7] Palmeri M, Mehnert J, Silk AW, et al. Real-world application of tumor mutational burden-high (TMB-high) and microsatellite instability (MSI) confirms their utility as immunotherapy biomarkers. ESMO Open. 2022 Feb;7(1):100336.

[8] Wheler JJ, Janku F, Naing A, et al. Cancer Therapy Directed by Comprehensive Genomic Profiling: A Single Center Study. Cancer Res. 2016 Jul 1;76(13):3690–701.

[9] Pleasance E, Bohm A, Williamson LM, et al. Whole-genome and transcriptome analysis enhances precision cancer treatment options. Ann Oncol. 2022 Sep;33(9):939–949.

[10] Simons M, Retel VP, Ramaekers BLT, et al. Early Cost Effectiveness of Whole-Genome Sequencing as a Clinical Diagnostic Test for Patients with Inoperable Stage IIIB,C/IV Non-squamous Non-small-Cell Lung Cancer. Pharmacoeconomics. 2021 Dec;39(12):1429–1442.

[11] Malone ER, Oliva M, Sabatini PJB, et al. Molecular profiling for precision cancer therapies. Genome Med. 2020 Jan 14;12(1):8.

[12] Nakagawa H, Fujita M. Whole genome sequencing analysis for cancer genomics and precision medicine. Cancer Sci. 2018 Mar;109(3):513–522.

[13] Alexandrov LB, Kim J, Haradhvala NJ, et al. The repertoire of mutational signatures in human cancer. Nature. 2020 Feb;578(7793):94-101.

[14] Shin HT, Choi YL, Yun JW, et al. Prevalence and detection of low-allele-fraction variants in clinical cancer samples. Nat Commun. 2017 Nov 9;8(1):1377.

[15] Faust GG, Hall IM. SAMBLASTER: fast duplicate marking and structural variant read extraction. Bioinformatics. 2014 Sep 1;30(17):2503–5.

[16] Van der Auwera GA, Carneiro MO, Hartl C, et al. From FastQ data to high confidence variant calls: the Genome Analysis Toolkit best practices pipeline. Curr Protoc Bioinformatics. 2013;43(1110):11 10 1-11 10 33.

[17] Cibulskis K, Lawrence MS, Carter SL, et al. Sensitive detection of somatic point mutations in impure and heterogeneous cancer samples. Nat Biotechnol. 2013 Mar;31(3):213–9.

[18] Chen X, Schulz-Trieglaff O, Shaw R, et al. Manta: rapid detection of structural variants and indels for germline and cancer sequencing applications. Bioinformatics. 2016 Apr 15;32(8):1220-2.

[19] McLaren W, Gil L, Hunt SE, et al. The Ensembl Variant Effect Predictor. Genome Biol. 2016 Jun 6;17(1):122.

[20] Dudley JC, Gurda GT, Tseng LH, et al. Tumor cellularity as a quality assurance measure for accurate clinical detection of BRAF mutations in melanoma. Mol Diagn Ther. 2014 Aug;18(4):409–18.

[21] Li MM, Datto M, Duncavage EJ, et al. Standards and Guidelines for the Interpretation and Reporting of Sequence Variants in Cancer: A Joint Consensus Recommendation of the Association for Molecular Pathology, American Society of Clinical Oncology, and College of American Pathologists. J Mol Diagn. 2017 Jan;19(1):4–23.

[22] Chan TA, Yarchoan M, Jaffee E, et al. Development of tumor mutation burden as an immunotherapy biomarker: utility for the oncology clinic. Ann Oncol. 2019 Jan 1;30(1):44–56.

[23] Alexandrov LB, Nik-Zainal S, Wedge DC, et al. Signatures of mutational processes in human cancer. Nature. 2013 Aug 22;500(7463):415-21.

[24] Burris HA, Saltz LB, Yu PP. Assessing the Value of Next-Generation Sequencing Tests in a Dynamic Environment. Am Soc Clin Oncol Educ Book. 2018 May 23;38:139–146.

[25] Zhong Y, Xu F, Wu J, et al. Application of Next Generation Sequencing in Laboratory Medicine. Ann Lab Med. 2021 Jan;41(1):25–43.

[26] Steuten L, Goulart B, Meropol NJ, et al. Cost Effectiveness of Multigene Panel Sequencing for Patients With Advanced Non-Small-Cell Lung Cancer. JCO Clin Cancer Inform. 2019 Jun;3:1–10.

[27] Hagemann IS, Devarakonda S, Lockwood CM, et al. Clinical next-generation sequencing in patients with non-small cell lung cancer. Cancer. 2015 Feb 15;121(4):631–9.

[28] Simons M, Ramaekers B, Peeters A, et al. Observed versus modelled lifetime overall survival of targeted therapies and immunotherapies for advanced non-small cell lung cancer patients - A systematic review. Crit Rev Oncol Hematol. 2020 Sep;153:103035.

[29] De Leeneer K, Hellemans J, De Schrijver J, et al. Massive parallel amplicon sequencing of the breast cancer genes BRCA1 and BRCA2: opportunities, challenges, and limitations. Hum Mutat. 2011 Mar;32(3):335–44.

[30] Bosdet IE, Docking TR, Butterfield YS, et al. A clinically validated diagnostic second-generation sequencing assay for detection of hereditary BRCA1 and BRCA2 mutations. J Mol Diagn. 2013 Nov;15(6):796–809.

[31] Zhao EY, Jones M, Jones SJM. Whole-Genome Sequencing in Cancer. Cold Spring Harb Perspect Med. 2019 Mar 1;9(3).

[32] Kim YN, Lee K, Park E, et al. Comprehensive genomic and immunohistochemical profiles and outcomes of immunotherapy in patients with recurrent or advanced cervical cancer. Front Oncol. 2023;13:1156973.

[33] Kim H, Kim R, Kim HR, et al. HER2 Aberrations as a Novel Marker in Advanced Biliary Tract Cancer. Front Oncol. 2022;12:834104.

[34] Kim S, Scheffler K, Halpern AL, et al. Strelka2: fast and accurate calling of germline and somatic variants. Nat Methods. 2018 Aug;15(8):591–594.

[35] Helleday T, Eshtad S, Nik-Zainal S. Mechanisms underlying mutational signatures in human cancers. Nat Rev Genet. 2014 Sep;15(9):585–98.

[36] Wen H, Feng Z, Ma Y, et al. Homologous recombination deficiency in diverse cancer types and its correlation with platinum chemotherapy efficiency in ovarian cancer. BMC Cancer. 2022 May 16;22(1):550.

[37] Tate JG, Bamford S, Jubb HC, et al. COSMIC: the Catalogue Of Somatic Mutations In Cancer. Nucleic Acids Res. 2019 Jan 8;47(D1):D941–D947.

[38] Pennell NA, Mutebi A, Zhou ZY, et al. Economic Impact of Next-Generation Sequencing Versus Single-Gene Testing to Detect Genomic Alterations in Metastatic Non-Small-Cell Lung Cancer Using a Decision Analytic Model. JCO Precis Oncol. 2019 Dec;3:1–9.

[39] Vanderpoel J, Stevens AL, Emond B, et al. Total cost of testing for genomic alterations associated with next-generation sequencing versus polymerase chain reaction testing strategies among patients with metastatic non-small cell lung cancer. J Med Econ. 2022 Jan-Dec;25(1):457-468.

